# EVIDENCE OF HIV/HCV COINFECTION AMONG PEOPLE LIVING WITH HIV/AIDS ATTENDING FEDERAL MEDICAL CENTRE, YENAGOA, NIGERIA

**DOI:** 10.1101/2023.03.08.23286986

**Authors:** IO Okonko, N Shaibu

## Abstract

Coinfection of hepatitis C (HCV) may compromise antiretroviral therapy (ART) in Nigeria. In this study, we evaluated the seroprevalence of HIV/HCV coinfection in people living with HIV/AIDs (PLWHA) receiving ART and associated factors. Patients were selected from HIV-1-infected patients enrolled in National HAART Cohort at Federal Medical Centre in Yenagoa, Nigeria. Following the manufacturer’s instructions, medical assessments and anti-HCV antibody serology were obtained for analysis with an ELISA kit (Dia. Pro). A total of 4 of 104 PLWHA tested were anti-HCV antibody positive (4.0%). HIV/HCV coinfections were higher in age groups ≥41 years (4.4%), males (7.0%), CD4 counts 350-499 cells/μl (7.1%) and PVL ≥1000 copies/ml (10.0%). CD4 counts and viral load were an indicator for HIV/HCV coinfections. Socio-demographic variables were not associated (p > 0.05) with HIV/HCV coinfection in univariate analysis; older PLWHA were more likely to be HCV-positive. Males were more prone to HIV/HCV coinfection than females. HIV status did seem to influence the predisposition to HCV infection, as an increase in susceptibility was observed with HIV-infected patients in Yenagoa, Nigeria. The high prevalence of HIV/HCV coinfection in PLWHA in Yenagoa receiving ART demands routine screening for viral hepatitis coinfection, intensive prevention of childhood HCV transmission, and modification of the management of HIV infection.

## 1. INTRODUCTION

Hepatitis C virus (HCV) is a significant cause of chronic hepatitis, liver cirrhosis, liver disease and hepatocellular carcinoma (HCC) worldwide (Cooke et al., 2013; Colpitts et al., 2020; Tsai et al., 2020; Hsu et al., 2022) and a common indication for liver transplantation (Patnaik & Tsai, 2022). Following infection, the majority of individuals will develop chronic hepatitis C that may subsequently lead to liver cirrhosis and cancer (Colpitts et al., 2020). Emerging evidence has indicated an association between HCV and atherosclerotic cardiovascular disease (Lee et al., 2019).

According to WHO (2023c), 290 000 persons died from hepatitis C in 2019, primarily from cirrhosis and hepatocellular carcinoma (primary liver cancer). Globally, it is estimated that 71 million people have chronic HCV infections (Polaris Observatory HCV Collaborators, 2017; Hsu et al., 2022). There is no vaccine to prevent HCV infection (Colpitts et al., 2020; WHO, 2023c); however, the development of safe and highly effective HCV therapy with direct-acting antiviral agents has revolutionized the management of HCV, liver transplant candidates and transplant recipients (Patnaik & Tsai, 2022).

About 38.4 million persons will live with the human immunodeficiency virus (HIV) globally in 2021, while about a quarter of this is in Africa (WHO, 2023a, b). The frequency of HIV is now becoming an epidemic in our world, especially in developing countries such as Nigeria. Usually, HIV infection has a long-term prognosis; some people get AIDS quickly and pass away within a few years if untreated (Okonko et al., 2020a, b). The presence of HCV further complicates this condition, as results from countries with highly active antiretroviral therapy (HAART) suggest liver diseases associated with HCV play a significant role in deteriorating health and increasing the mortality rate in persons living with HIV (Konopnicki et al., 2005; Okonko et al., 2020a, b, WHO, 2020). Another significant obstacle is coinfection since it slows the progression of the illness to AIDS (Okonko et al., 2020a, b).

Human Immunodeficiency Virus (HIV), hepatitis B virus (HBV), and hepatitis C virus (HCV) are the three most common chronic viral infections of public health importance with significant socio-economic impact worldwide (Muriuki et al., 2013; Esan et al., 2014; Lawal et al., 2020). Both HBV and HCV infections cause acute and chronic liver infections with the potential for liver cirrhosis and hepatocellular carcinoma (HCC) (Cooper et al., 2009; Abera et al., 2014; Lawal et al., 2020). Hepatitis C leads to chronic infection in 60.0–80.0% of patients after acute infection (Sadoh et al., 2011; Ejeliogu et al., 2014; Lawal et al., 2020), while perinatal HBV infection is associated with a 90.0% risk of chronicity compared with a risk of less than 5.0% among adults with intact immunity (Paganelli et al., 2012; Lawal et al., 2020).

Globally, over 350 million and 170 million people are chronically infected with HBV and HCV, respectively (Nel et al., 2012; Abera et al., 2014; Lawal et al., 2020). In Africa, about 100 million individuals are estimated to be infected with HBV or HCV (Lemoine et al., 2015; Lawal et al., 2020). Furthermore, 80.0% of liver cirrhosis and HCC cases in Africa are caused by HBV and HCV infections, with HBV being the primary contributor to end-stage liver disease (Lemoine et al., 2015; Lawal et al., 2020).

Although varied, the prevalence of HCV/HIV coinfection is lower than that of HBV/HIV coinfection worldwide. In a large group of children who were perinatally HIV-positive and were part of a long-term follow-up program, the prevalence of HCV infection in the United States was 1.5%, compared to a rate of 9.6% in China (Schuval et al., 2004; Zhou et al., 2010; Lawal et al., 2020). However, several African nations, such as Tanzania, where a prevalence of 13.8% has been recorded, and Ethiopia, where the prevalence of anti-HCV ranged from 0.0% to 22.0%, have seen a higher incidence of this coinfection (Deress et al., 2021) On the other side, Cote d’ Ivoire in West Africa recorded a prevalence rate of 0.0% (Telatela et al., 2007; Rouet et al., 2008; Riou et al., 2016; Lawal et al., 2020) and in Ivory Coast and Kenya (Puoti et al., 2008; Rouet et al., 2008). Similarly, studies in Port Harcourt, Nigeria, and East Africa found 0.0% prevalence rates for HIV/HCV coinfection (Chakraborty et al., 2003; Okonko et al., 2014; Lawal et al., 2020; Cookey et al., 2021). Lagos, Nigeria, reported a 0.5% prevalence incidence of HIV/HCV coinfection (Lawal et al., 2020).

Its endemicity has long been validated in Nigeria, but there is less information about its epidemiology (Riou et al., 2016; Abeni et al., 2020; Okonko & Ernest Nwagwu, 2023). There is an established relationship between HIV positivity and the progression of HCV infection; dual positivity with HIV influences the clinical outcome of HCV infection (Villano et al., 1999; Thomas et al., 2000; Sulkowski & Thomas, 2003; Ogbodo et al., 2015; Okonko et al., 2022). The alarming statistics have made it crucial that studies be also done to ascertain the HIV/HCV coinfection prevalence in the country and host factors which may influence the coinfection (Cookey et al., 2021).

We know very little research on HCV, coinfections among HIV-positive individuals, and related risk factors in Bayelsa State, Nigeria. In order to establish a baseline of knowledge, this study was carried out to ascertain the serological evidence, prevalence, and coinfection rates among HIV-positive individuals in Yenagoa, Nigeria.

## 2. MATERIAL AND METHODS

### 2.1. Study Area

The Federal Medical Center in Yenagoa, Bayelsa State, Nigeria, was the site of the study. This hospital is one of the primary treatment centres in Bayelsa State, Southern Nigeria, for people living with HIV and AIDS (PLWHA). Yenagoa is a Local Government area and city of Bayelsa State, Nigeria. It is located in the southern part of the country. The LGA has an area of 706 km^2^ and a population of over 352,285 as of 2006. The Ijaw form the majority of the state. Bayelsa is situated in an area of swamps, mangroves, and tropical rainforests, and it is located in the core of the Niger Delta region. Bayelsa state was formed in 1996 by Rivers State, making it one of the newest in the federation. The state bothers River State, of which it was formerly part of and Delta state. The state is the smallest in Nigeria by population as of the 2006 Census and one of the smallest by area. Bayelsa State has a riverine and estuarine setting, with bodies of water within the state preventing the development of significant road infrastructure. Petroleum is the primary sector of the Bayelsa economy.

### 2.2. Study Design

The current investigation, which aims to ascertain the prevalence of HIV/HCV coinfections among people living with HIV and AIDS (PLWHA) attending Federal Medical Centre, Yenagoa, Bayelsa State, Nigeria, utilized a hospital-based cross-sectional study design. The method for this study consists of informed consent and blood withdrawal by venipuncture. Screening for HIV/HCV coinfections, clinical evaluation and recording of demographic information such as the age of the participants, sex, marital status, educational background, occupation, and use of ART.

### 2.3. Ethics statement

Administrative approval for this study was obtained from the management of the Federal Medical Centre, Yenagoa, Nigeria. Ethical considerations and approval for the study were obtained from the University of Port Harcourt Research Ethics Committee following the ethics for research involving human subjects. Before samples were obtained and analyzed, all subjects provided their informed consent. General information regarding the nature of the study and its objectives was explained to participants, who were also informed of the right of refusal to participate in the study or to withdraw at any time without jeopardizing their right to access other health services. Identification codes were used instead of names, and the information collected was kept confidential. This study followed the World Medical Association’s (WMA) Declaration of Helsinki, which sets forth the standards for medical research involving identified human/animal subjects, human subjects, and animal subjects.

### 2.4. Study population

Here, 104 PLWHA positively confirmed patients attending the HIV outpatient clinic of Federal Medical Centre, Yenagoa, Nigeria, who willingly gave informed consent and volunteered to have their blood samples examined were recruited into the study. Patients attended the clinic for routine check-ups, collection of medications or other medical complaints. Socio-demographic data of the patients were collected. Necessary demographics (age, sex, marital status, educational background and occupations) and clinical and epidemiological data of each patient were obtained using a well-structured questionnaire. Health personnel conducted the interview and entered the data according to the pre-structured questionnaire. All PLWHA positively confirmed patients were eligible for the study. HIV-infected patients with duplicate records or missing information about their socio-demographic data were excluded from the study. However, PLWHA, who were accurately recorded in the registration book, was included.

### 2.5. Sample collection

The method of sample collection employed was the vein puncture technique. A soft tourniquet was fastened to the upper hand of the patient. The punctured site was cleansed with methylated spirit, and the vein was punctured with a 3ml syringe. After sufficient blood collection, the tourniquet is released, and the needle is removed immediately. About 3 ml of venipuncture blood was collected in EDTA BA Vacutainer TM anti-coagulant tubes (BD, Franklin Lakes, USA), labelled with each patient’s details. Plasma specimens were separated by centrifugation at 3 000 rpm (revolution per minute) for 5 min. The plasma utilized for the laboratory tests was kept at -20°C.

### 2.6. Serological analysis of HCV Antibody

Blood samples of HIV-positive patients were taken using the venipuncture technique. In the Virology & Genomics Research Unit, Department of Microbiology, University of Port Harcourt, Nigeria, plasma was examined for anti-HCV antibodies using an enzyme-linked immunosorbent assay using a commercial kit (DIA.PRO Diagnostic Bioprobes, Milano, Italy) (ELISA). Laboratory testing was carried out according to the manufacturer’s instructions, and all tests were done utilizing quality controls according to standard operating procedures. A microplate reader for an ELISA was used to read the optical signals produced in the microwells at 450 nm (Model ELx808i, BioTek Instruments, USA). The formula for calculating the cut-off OD450nm (OD of negative control plus 0.350), which we utilized as a threshold for differentiating between reactive and non-reactive blood samples, was provided by the manufacturer of the ELISA kit. In order to determine the following cut-off formulation, the test results were derived using the mean OD450nm value of the negative control (NC) and a mathematical calculation: NC + 0.350 is cut-off (CO). Samples were deemed non-reactive for antibodies specific to the HCV antigens present if their OD450nm was below the cut-off value. Samples were deemed to have antibodies specific to the HCV antigens present if their OD450nm was more significant than the Cut-Off value. According to the manufacturer’s instructions, the results were interpreted. The values obtained were used to interpret the results: S/CO 0.9 negative, 0.9-1.1 equivocal, and >1.1 positives. The results were calculated as the ratio of the sample OD450nm and the cut-off value.

### 2.7. CD4 and Viral Load Analysis

Blood samples were analyzed for CD4+ T cell estimates by flow cytometry and viral load by Abbott Real-Time protocol.

### 2.8. Data analysis

Data were analyzed using SPSS version 20.0 (SPSS Inc. Chicago, IL, USA). HIV/HCV coinfections among PLWHA were compared to CD4+ T cell count, viral loads, socio-demographic variables and ART using Pearson’s chi-square (χ ^2^) or Fisher’s exact test, where appropriate. Statistical significance for all analyses was determined at a 5% significance level.

## 3. RESULTS

### 3.1. Study Population Characteristics

Here, we describe the result of the serological analysis of 104 HIV-positive patients attending the antiretroviral clinic at the Federal Medical Centre, Yenagoa. The participants’ age, sex, marital status, educational attainment, occupations, CD4 counts, and plasma viral loads were stratified, and all of the samples underwent serological analysis. Table 1 highlights the characteristics of the study group.

**Table 1:** Patients Characteristics.

| **Variables** | **Categories** | **No. Tested** | **Percentage (%)** |
| --- | --- | --- | --- |
| **Age groups (Years)** | 8-20 | 8 | 7.7 |
|  | 21-40 | 51 | 49.0 |
|  | 41 & above | 45 | 43.3 |
| **Sex** | Females | 75 | 72.1 |
|  | Males | 29 | 27.9 |
| **Marital Status** | Singles | 43 | 41.4 |
|  | Married | 56 | 53.9 |
|  | Divorced | 5 | 4.8 |
| **Educational Background** | Primary | 17 | 16.4 |
|  | Secondary | 43 | 41.4 |
|  | Tertiary | 42 | 40.4 |
|  | None | 2 | 1.9 |
| **Occupations** | Self-Employed | 27 | 26.0 |
|  | Unemployed | 10 | 9.6 |
|  | Business/Trader | 29 | 28.0 |
|  | Students | 23 | 22.1 |
|  | Artisans | 7 | 6.7 |
|  | Civil Servants | 8 | 7.7 |
| **CD4 Counts (Cells/μl)** | <200 | 54 | 52.0 |
|  | 200-349 | 10 | 9.6 |
|  | 350-499 | 14 | 13.5 |
|  | 500 & above | 26 | 25.0 |
| **Viral Loads (Copies/ml)** | <20 | 53 | 51.0 |
|  | 20-999 | 41 | 39.4 |
|  | 1000 & above | 10 | 9.6 |
| **Total** |  | **104** | **100.0** |

### 3.2. HIV/HCV Coinfection

Overall, HIV/HCV coinfection was determined to be 4.0% (Figure 1).

**Figure 1:**
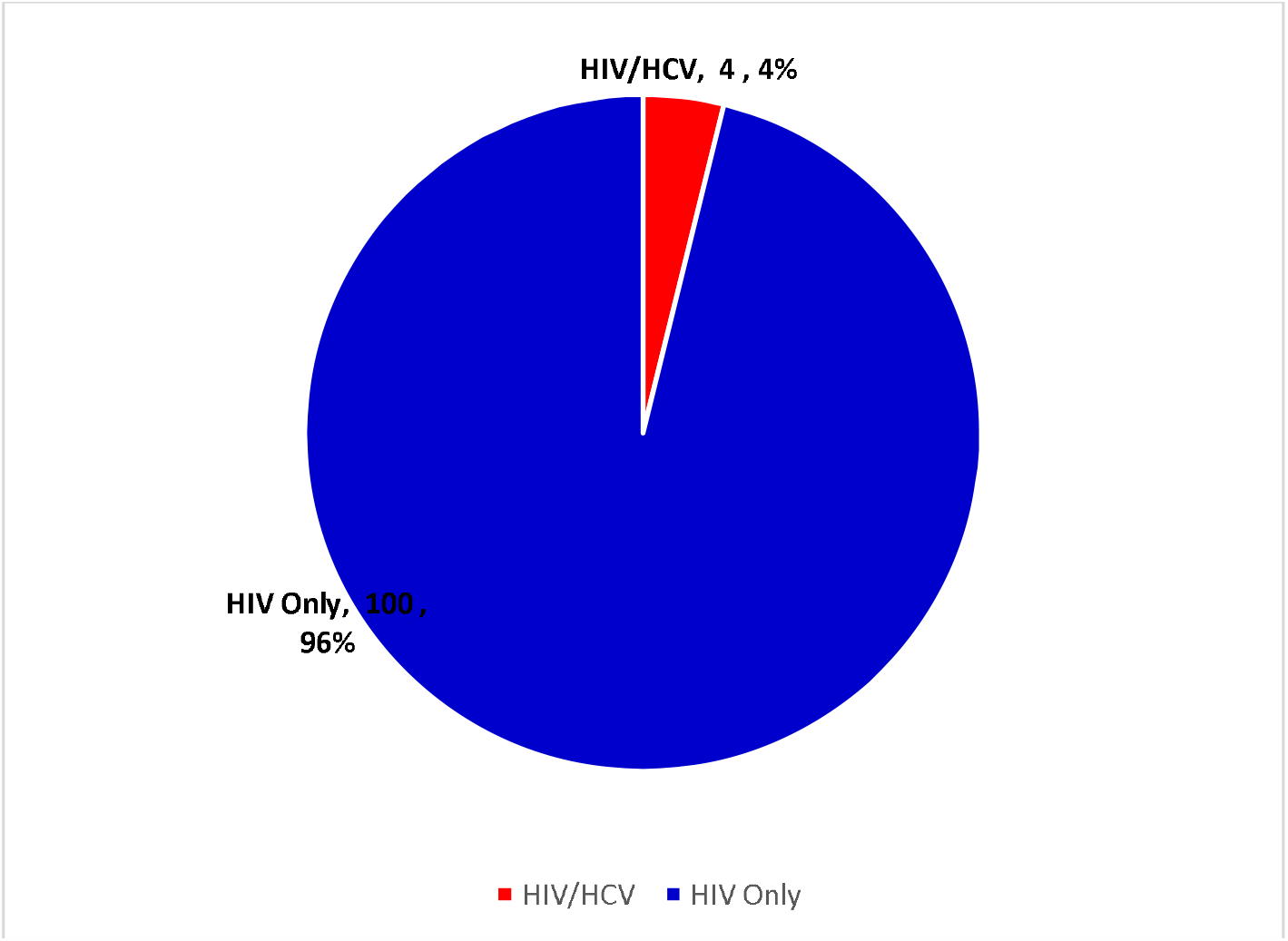
Overall HIV/HCV coinfection.

### 3.3. Age-specific HIV/HCV coinfection

Higher HIV/HCV coinfection also occurred among age groups <u>≥</u>41 years (4.4%) than 21–40 years (4.0%) and <u>≤</u>20 years with 0.0% (Figure 2). These differences were not statistically associated (p = 0.83).

**Figure 2:**
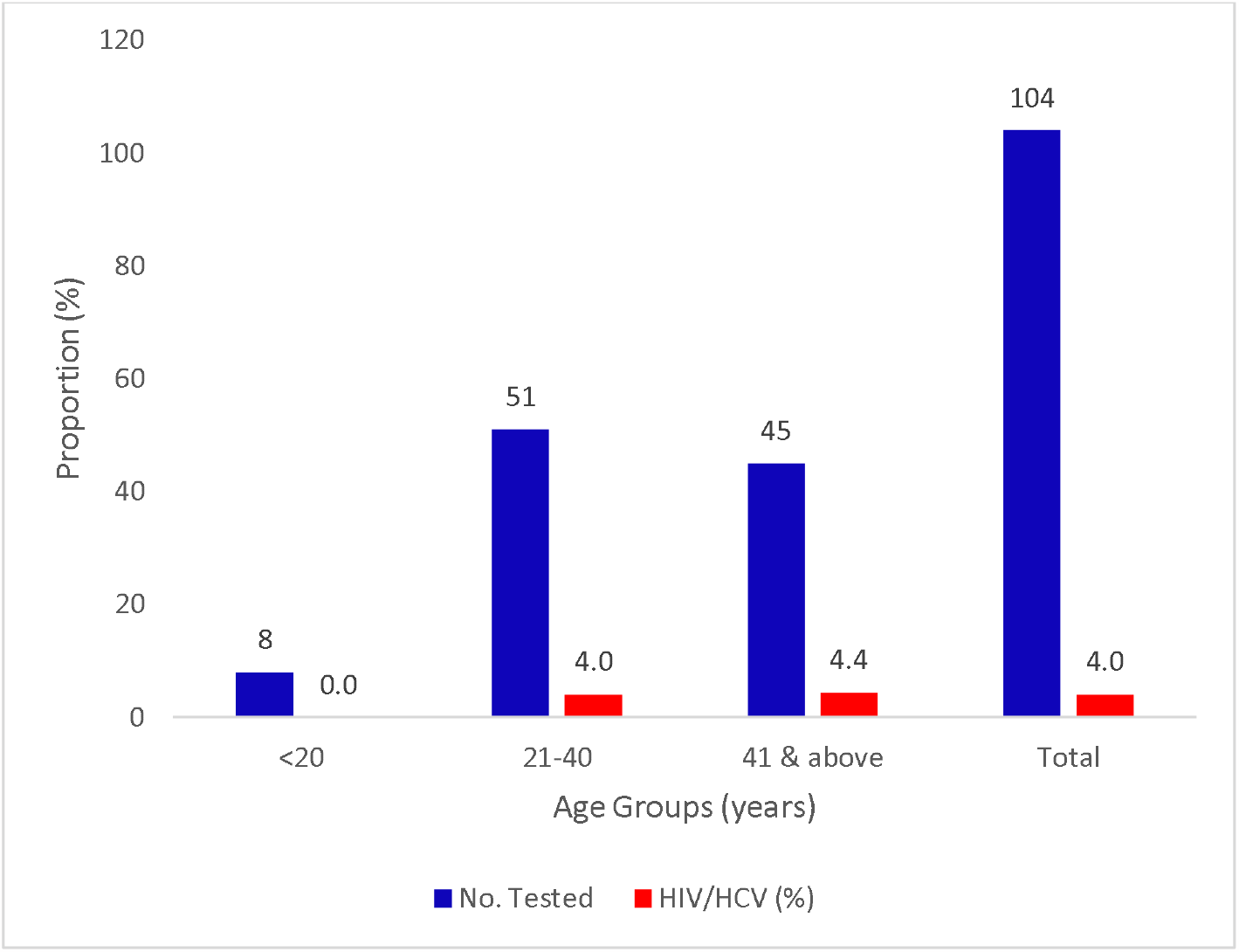
HIV/HCV coinfections with Age.

### 3.4. Sex-specific HIV/HCV coinfection

Higher HIV/HCV coinfection was observed among males (7.0%) than in females (3.0%) (Figure 3). No significant association existed between HIV/HBV coinfection and sex (p = 0.31).

**Figure 3:**
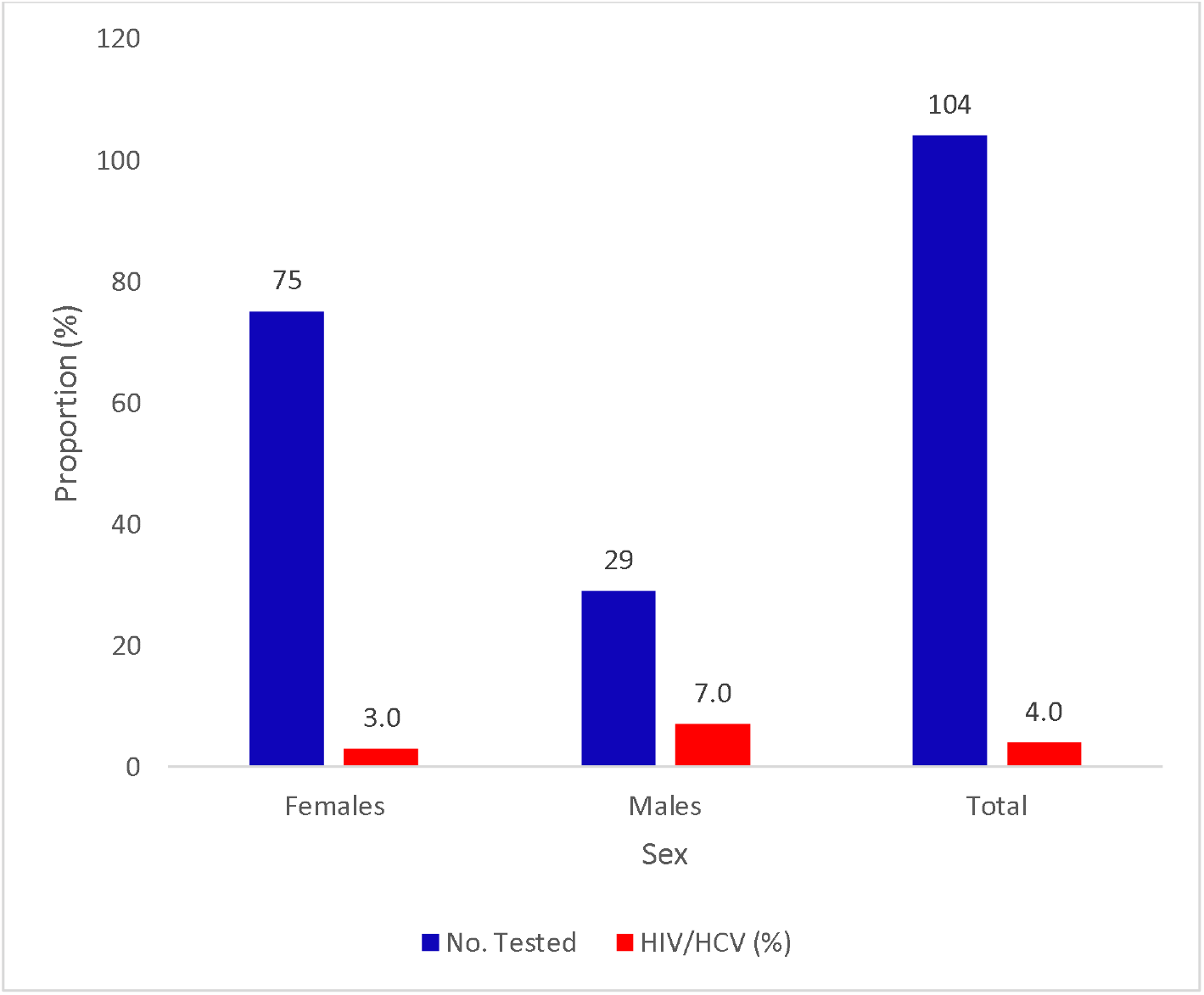
HIV/HCV coinfections with Sex.

### 3.5. Marital Status-specific HIV/HCV coinfection

Higher HIV/HCV coinfection occurred among singles (5.0%) than the married (4.0%) and divorced (0.0%) (Figure 4). No significant association existed between HIV/HCV coinfection and marital status (p = 0.87).

**Figure 4:**
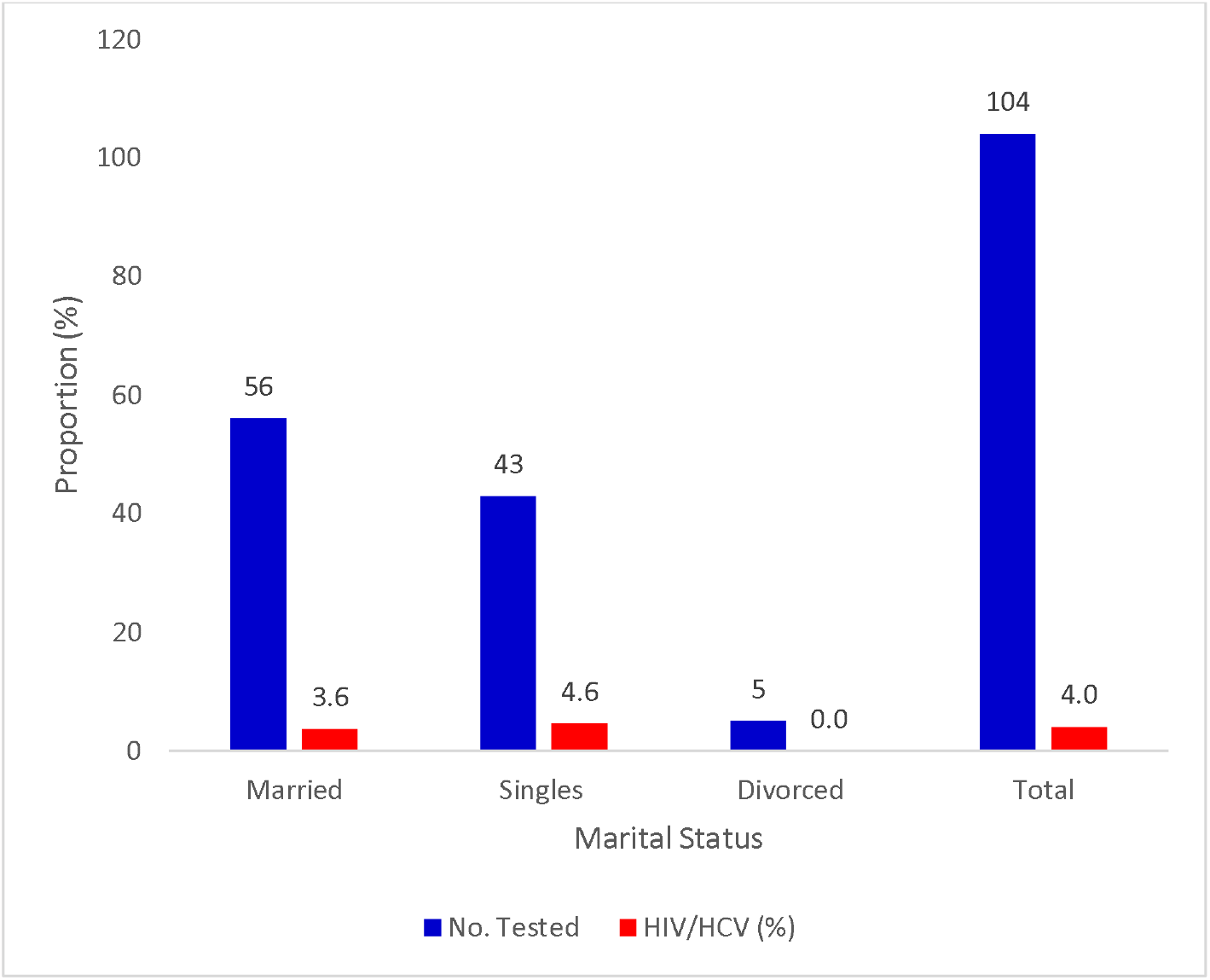
HIV/HCV coinfections with Marital Status.

### 3.6. Educational Background-specific HIV/HCV coinfection

Higher HIV/HCV coinfection was observed among those with tertiary educational backgrounds (5.0%) than those with secondary education (4.6%) and primary education with 0.0% (Figure 5). No significant association existed between HIV/HCV coinfection and educational background (p = 0.82).

**Figure 5:**
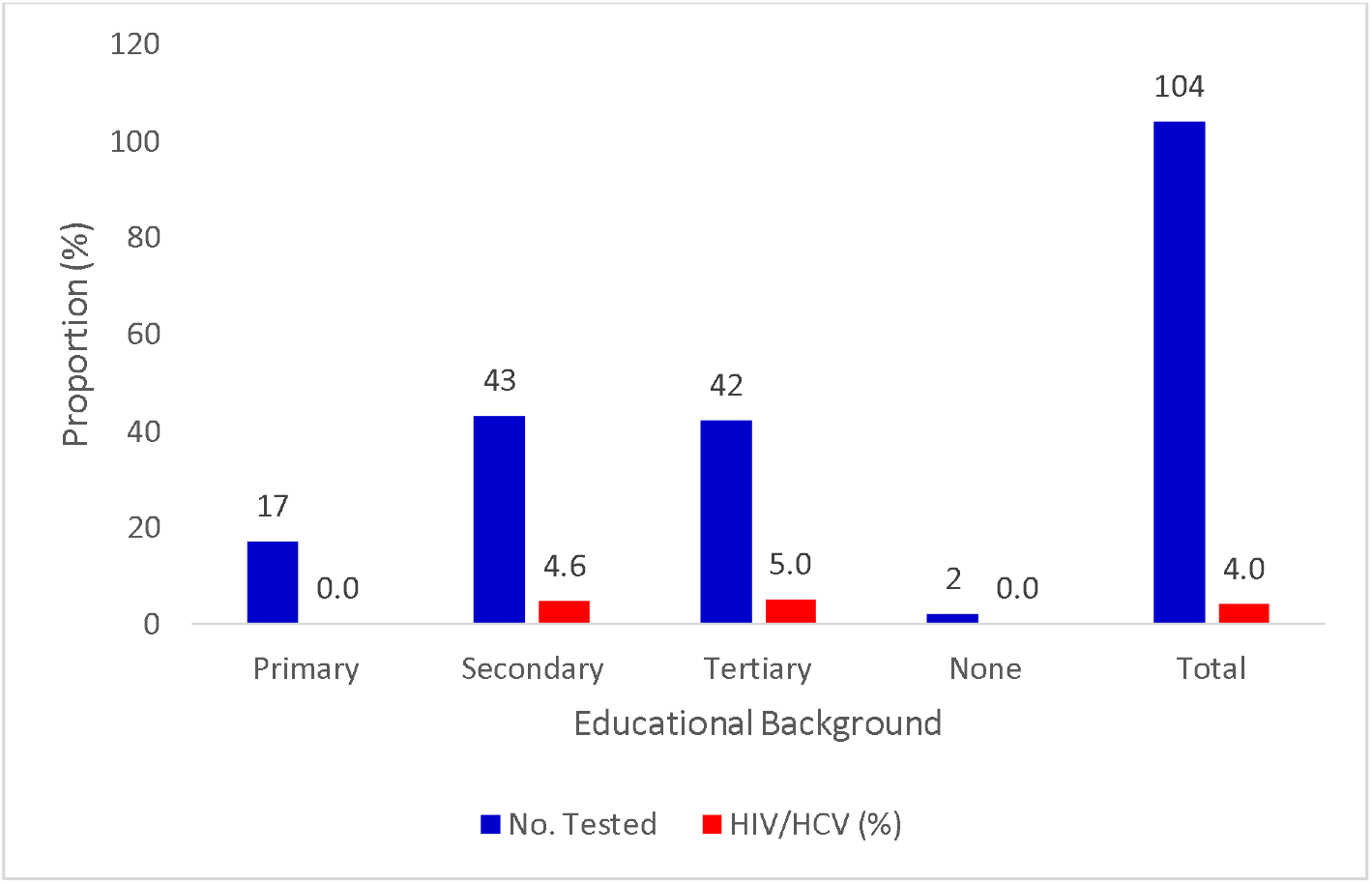
HIV/HCV coinfections with Educational Background.

### 3.7. Occupation-specific HIV/HCV coinfection

Higher HIV/HBV coinfection occurred among self-employed (7.4%) than business/traders (7.0%) and other occupations with 0.0% (Figure 6). No significant association existed between HIV/HCV coinfection and occupations (p = 0.61).

**Figure 6:**
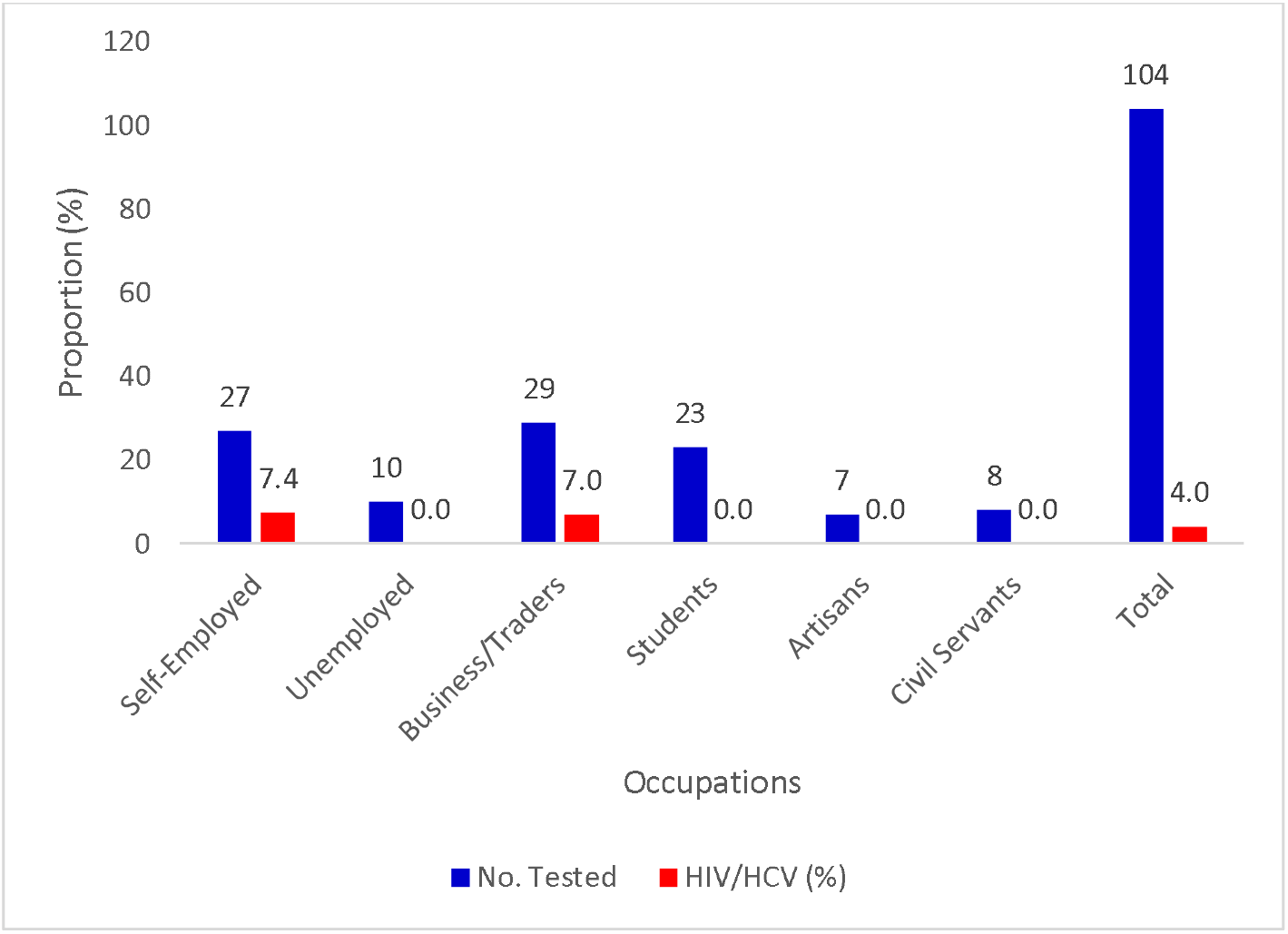
HIV/HCV coinfections with Occupation.

### 3.8. CD4 Counts-Related Specific HIV/HCV coinfection

In terms of CD4 counts, higher HIV/HCV coinfection (7.1%) was observed for participants with CD4 counts 350-499 cells/μl than <200 cells/μl (5.6%) and <u>≥</u>500 cells/μl (3.8%) while 200-349500 cells/μl had the least, 0.0% (Figure 7). No significant association existed between HIV/HCV coinfection and CD4 counts (p = 0.85).

**Figure 7:**
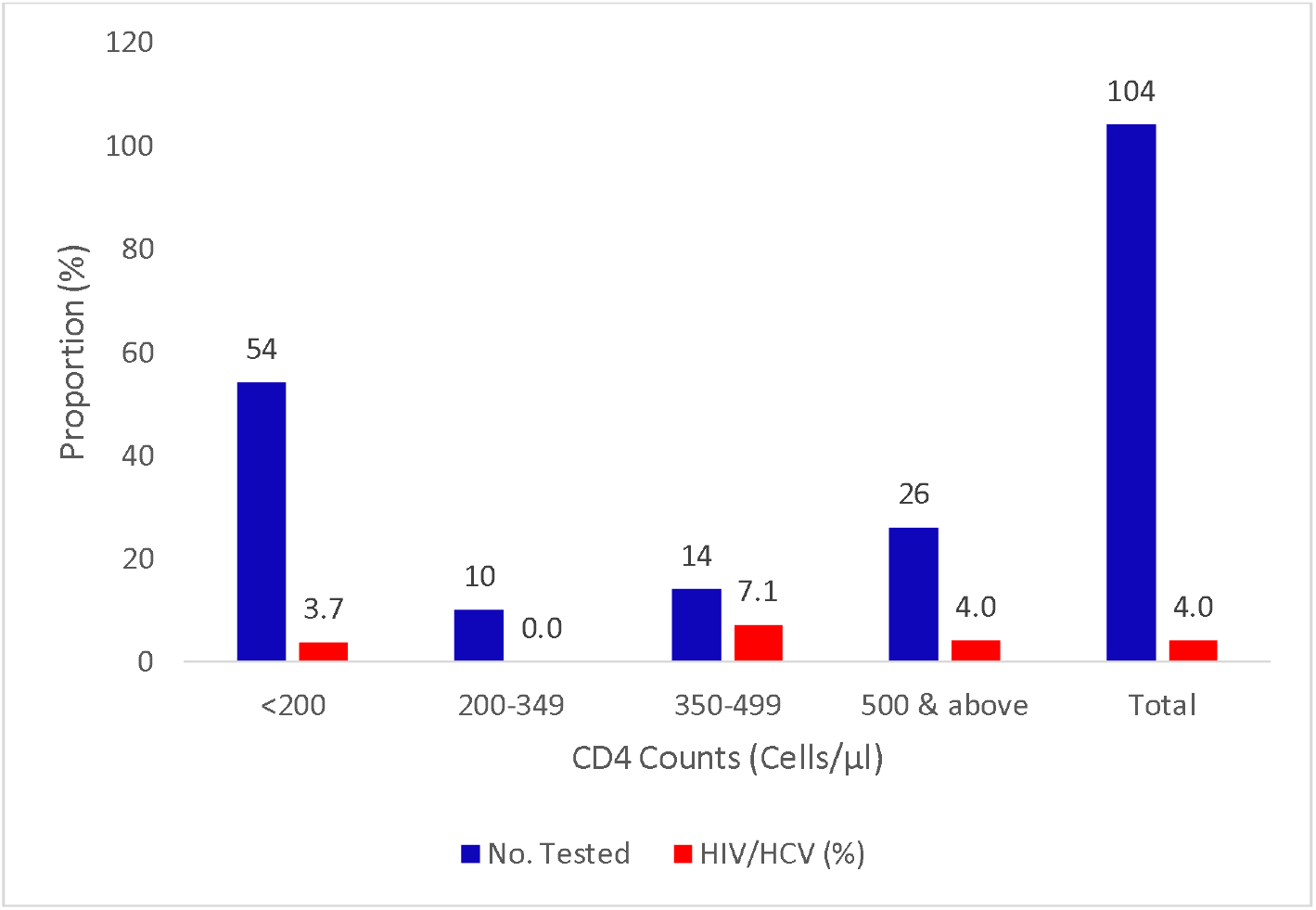
HIV/HCV coinfections with CD4 Counts.

### 3.9. Viral Loads-Related Specific HIV/HCV coinfection

In terms of viral loads, higher HIV/HCV (10.0%) occurred among participants that had <u>≥</u>1000 copies/ml than those with <20 copies/ml (3.8%) and 20-999 copies/ml with 2.4% (Figure 8). No significant association existed between HIV/HCV coinfection and viral loads (p = 0.54).

**Figure 8:**
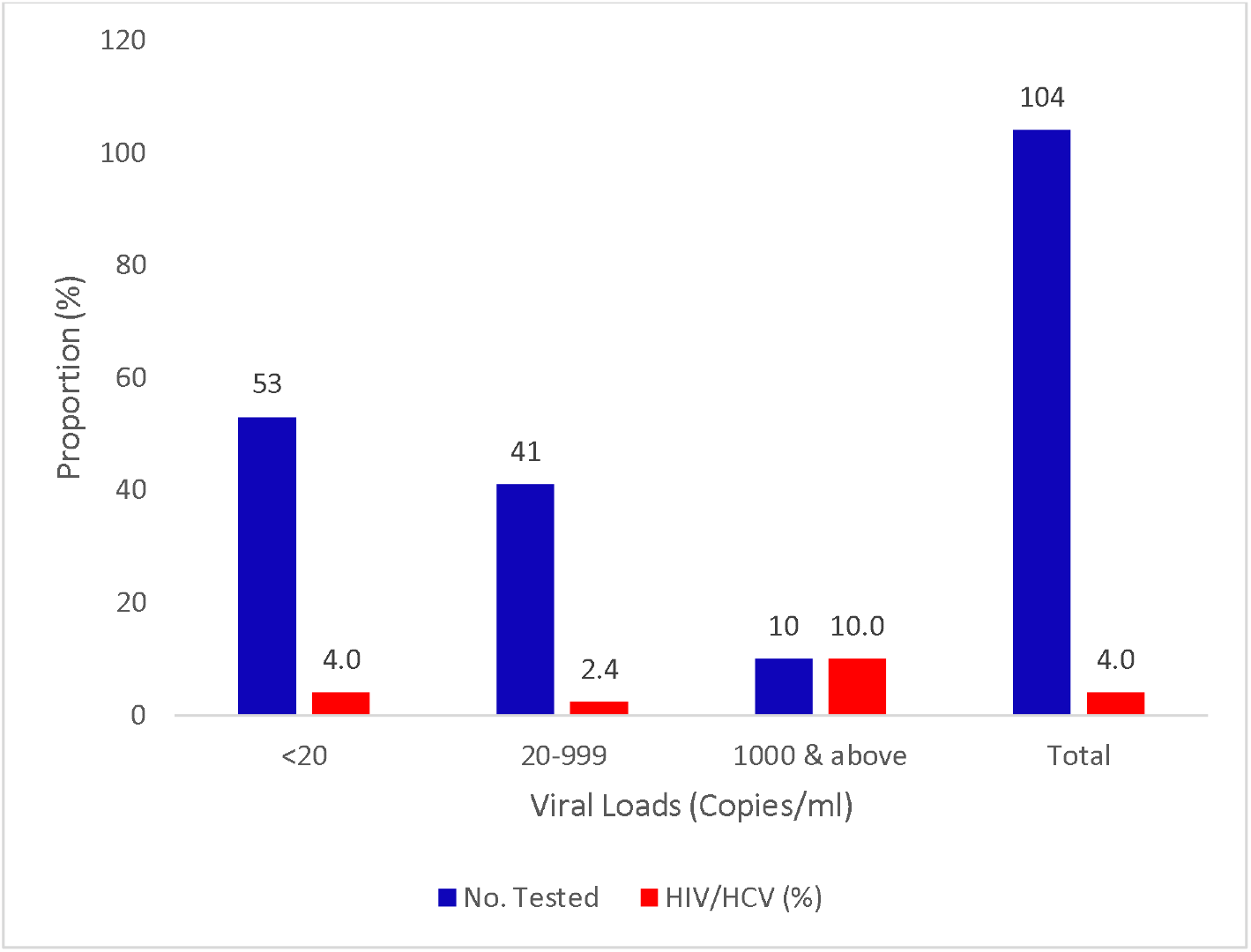
HIV/HCV coinfections with Viral Loads.

## 4. DISCUSSION

As much as 2%–3% of the world’s population may be infected with the hepatitis C virus (HCV), making it a significant worldwide health concern (Alter et al., 2007; Mohd Hanafiah et al., 2013; Benova et al., 2014). Because it persists in about 85.0% of infected people, HCV infection is clinically significant because it presents a serious risk of chronic liver damage (Omolade & Adeyemi, 2018; Okonko & Ernest Nwagwu, 2023). Antiretroviral therapy (ART) in Nigeria could be compromised by coinfection with the hepatitis C virus (HCV). To establish a baseline for future research, this study examined the serological prevalence of HCV and coinfections among people with HIV/AIDS (PLWHA) in Yenagoa, Nigeria. HCV had a 4.0% coinfection prevalence rate among the 104 PLWHA. This result implies little evidence of HCV and HIV coinfection among research participants.

The 4.0% coinfection rate with HCV contradicts the 15.0% and 23.5% reported by Ogbodo et al. (2015) and Ogwu-Richard et al. (2015) in Nigeria. It also contradicts the findings of Okonko et al. (2014) and Cookey et al. (2021), who determined that 0.0% of Port Harcourt, Nigeria, residents have HCV antibodies. Elsewhere, it is comparable to Tsai et al. (2015), who reported a 4.7% HIV/HCV coinfection in Southern Taiwan and Ionita et al. (2017), who reported 4.4% in Nepal. Moreover, it is comparable to China’s 4.9% (Zhou et al., 2010).

Contrarily, the rate of HIV/HCV coinfection in this study is higher than that of studies done in Ethiopia, which revealed a 1.3% rate (Balew et al., 2014). This 4.0% is also higher than the 0.5% for HIV/HCV coinfection among HIV-infected and HIV-naive children in Lagos (Lawal et al., 2020), the 1.7% in Northcentral, Nigeria (Durowaye et al., 2014), the 1.5% in the USA (Schuval et al., 2004), 1.0% was recorded by Aaron et al. (2021) in Port Harcourt, Nigeria, the 0.16% prevalence in the southeast of Nigeria (Ikeako et al. (2014), 0.5% in Anyigba (Omatola et al., 2019), 0.7% in the southeast (Diwe et al., 2013), 0.8% in Abuja (Agboghoroma & Ukaire, 2020), 0.8% in Osogbo (Oluremi et al. (2021), 0.9% reported by Onyekwere and Hameed (2015) and 2.0% in Ethiopia (Deress et al., 2021).

The prevalence of HIV/HCV coinfection in this study was 4.0%, lower than the 5.8%, 5.8%, and 6.02% found in Southeast and South-South, Nigeria (Ikpeme, 2013; Nwolisa et al., 2013). This 4.0% is also lower than the 5.5% in Kumasi, Ghana (Boateng et al., 2019), 7.0% and 7.5% in Lagos, Nigeria (Lawal et al., 2020), 8.1% in Ondo State, Nigeria (Ogundele et al., 2017), 9.6% in China (Zhou et al., 2010) and 22.5% in Port Harcourt, Nigeria (Okonko et al., 2022). It is also lower than the 5.8% and 10.8% recorded by Benova et al. (2014), the 5.2% and 5.0% in Benin and Jos, respectively (Sadoh et al., 2011; Ejeliogu et al., 2014). Compared to China’s 62.4% and 50.2% claims, the 4.0% is significantly lower (Chen et al., 2013). The mixed population could explain these results that some authors studied in Nigeria, where teenagers were more likely to have HIV/HCV coinfections (Sadoh et al., 2011). The people analyzed were all HAART-experienced, which may also explain the low prevalence of coinfection reported in the cohort in contrast to Ejeliogu et al. (2014); all their study participants were HAART naïve.

According to prevalence reports from Nigeria and other countries (Oni & Harrison, 1996; Ogboghodo et al., 2009; Khaderi et al., 2014; Eke et al., 2015; Fox et al., 2015; O’Learya et al., 2017), the prevalence rate of 4.0% for HIV/HCV in the current study is not comparable to prevalence reports from those countries. This study further supports the low frequency of HCV in children and the general population. In contrast to the prevalence in this study, greater prevalence rates of 10.0% in Northern Nigeria and 30.30% in Plateau State, Nigeria, were reported by Isa et al. (2015) and Odjimogho et al. (2018), respectively. This observation is also related to the variation in the distribution of HCV infection and the fact that the sample size in the prior study was slightly similar. Similar transmission routes for viruses in the study participants and the restricted availability of health information regarding the spread and control of HCV infection in the study region could be potential causes of the disparities in this investigation.

The coinfection rate of HIV/HCV in this study was significantly higher among older participants, which differs from a study reported by Ogbodo et al. (2015) in Ughelli, Delta State, Nigeria and Abeni et al. (2020) in Port Harcourt, Nigeria. However, it corroborates our recent study in Port Harcourt, Nigeria (Okonko et al., 2022) and Ogbakiri, Emohua LGA, Rivers State, Nigeria (Okonko & Ernest Nwagwu, 2023). It also supports Tessema et al.’s (2010) study in Plateau State, Nigeria (Odjimogho et al., 2018). It also supported the observation made in a study in China (Chen et al., 2013) and Taiwan (Tsai et al., 2021). This study indicates that the elderly are more susceptible to HCV/HIV coinfection than other age groups. This result might be the weakened immune system of the elderly and end-stage liver disease complications. Therefore, it can be assumed that age is a very significant independent predictor of HIV/HV coinfection.

The current study recorded a higher prevalence among males (7.0%) than females (3.0%). Contributory factors may be attributed to socio-economical, hormonal, or immunological factors. Sexual activities and societal freedom can also influence the prevalence rate of HIV/HCV coinfection in males. This observation is similar to Gupta et al. (2018), who recorded that males were found to be HCV positive in greater percentage than females, which may be assumed that in India, males look for healthcare services earlier than females. Rao et al. (2015) and Odjimogho et al. (2018) have reported findings contrary to this observation. This high infection rate in males than in females may be due to the spontaneous clearance of acute infection in females. The reason for this clearance has been attributed occurrence of certain genetic factors. Other researchers from other parts of Nigeria have verified this demographic tendency (Egah et al., 2004; Buseri et al., 2009; Udeze et al., 2009; Okonko et al., 2012; Kassim et al., 2012; Okocha et al., 2015; Ionita et al., 2017; Okoroiwu et al., 2018; Abeni et al., 2020) and a study in China (Chen et al., 2013). However, this observation differs from other studies in Nigeria (Lesi et al., 2007; Udeze et al., 2011; Balogun et al., 2012; Jemilohun et al., 2014; Ogbodo et al., 2015; Okonko et al., 2022; Okonko & Ernest Nwagwu, 2023).

The current study showed that HCV prevalence was highest in singles. This observation is discordant with the reports of Umumararungu et al. (2017), who documented a higher prevalence among married and divorced/widowed people than singles. Our observation supports previous studies in Nigeria, which observed higher rates among singles than married (Abeni et al., 2020; Okonko et al., 2022; Okonko & Ernest Nwagwu, 2023). This high prevalence may imply that transmission of HEV via sex is essential. To break the cycle of HCV transmission, it is highly recommended that screening among sexually active individuals be performed.

Furthermore, a higher HIV/HCV coinfection occurred among those with a tertiary educational background than those with secondary and primary education. This observation supported a prior study in Nigeria (Okonko et al., 2022) and disputed the findings of other authors (Onyekwere et al., 2016; Ionita et al., 2017; Okonko & Ernest Nwagwu, 2023).

Concerning occupation, the positivity of HCV/HIV occurred more in self-employed (7.4%) and business/traders (7.0%) than in other occupational groups (0.0%). It can be assumed that these individuals reside in rural areas where there is a lack of public awareness/enlightenment about HIV/HCV coinfection. Logical reasoning could also be that, although uncommon, these individuals can become infected when healthcare professionals do not follow the proper steps to prevent the spread of blood-borne infections, share personal items, and have sex with an infected person. It can be assumed that most of these fore mentioned individuals might be traditional practitioners. Unsafe injections by traditional practitioners could be that these individuals are observed to have little or low educational attainment resulting in ignorance and naiveness.

Regarding CD4 counts, study participants who had HIV/HCV coinfection in this study have higher CD4 count (350-499 cells//μl), which is incomparable with the studies conducted in South Africa and Southern Nigeria (Lodenyo et al., 2000; Olufemi et al., 2009). These contentious findings may result from the study’s viral hepatitis or may be related to individual participant immune status disparities. In individuals with HIV/HCV coinfections, there may be high HIV and HCV viral replication that may further impair the patient’s immune system.

Regarding viral loads, higher HIV/HCV occurred among participants with viral load ≥1000 copies/ml than those with <20 copies/ml and 20-999 copies/ml. Contrary to our expectations, immunity was boosted, as indicated by increased CD4+ T-cell count in the HIV/HCV group compared to that of the HIV mono-infection group. Even the rate was twice as high as the HIV/HCV group. However, a similar viral load was recorded in the HIV/HCV and HIV mono-infection, respectively (Boateng et al., 2019). One school of thought could be that HCV inhibits or reduces CD4+ T-cells clearance by HIV (Boateng et al., 2019). Another line of reasoning is that HCV, as previously documented, promoted CD4+ T-cell activation (BenMarzouk-Hidalgo et al., 2016; Boateng et al., 2019).

Many demographic, virological, clinical, and lifestyle factors are linked to the natural course of HCV infection. These factors differ significantly across populations, locations, and time (WHO, 2017; Yu et al., 2021).

In this study, none of the socio-demographic characteristics of these participants was significantly associated (p > 0.05) with HIV/HCV coinfections.

## 5. CONCLUSION

In Yenagoa, Nigeria, HCV coinfections among people with HIV and AIDS (PLWHA) have been further validated by the current investigation. The total prevalence of HIV/HCV coinfections in this study was 4.0%. Viral load and CD4 counts served as indicators for HIV/HCV coinfections. Whereas their female counterparts showed a larger propensity to HIV infection alone, males had a higher propensity for HIV/HCV coinfection. Notwithstanding the low rates of HCV coinfection seen in this study, routine testing for HCV indicators should be done to lower morbidity and death in this group. More research is advised to comprehend ART’s effect on HCV in PLWHA fully.

## Data Availability

All data produced in the present work are contained in the manuscript.

## Compliance with ethical standards

## Acknowledgements

The authors would like to acknowledge the support obtained from the management and staff of Federal Medical Centre, Yenagoa, Bayelsa State, Nigeria, Nigeria, during the enrollment and collection of samples used in this study. The authors are grateful to the participants for their willingness to be part of the study and to Dr T. I. Cookey for the statistical analysis.

## Statement of ethical approval

The Federal Medical Centre in Yenagoa, Nigeria, offered their administrative authorization for this study. All authors affirm that the University of Port Harcourt Research Ethics committee has reviewed and approved all experiments. As a result, the study is conducted in accordance with the ethical guidelines outlined in the 1964 Declaration of Helsinki.

## Statement of informed consent

“All authors declare that all individuals participating in the study gave their free and informed permission.”

